# Shifting Geographical Transmission Patterns: Characterizing the 2023 Fatal Dengue Outbreak in Bangladesh

**DOI:** 10.1101/2024.03.24.24304789

**Authors:** Mohammad Nayeem Hasan, Mahbubur Rahman, Meraj Uddin, Shah Ali Akbar Ashrafi, Kazi Mizanur Rahman, Kishor Kumar Paul, Mohammad Ferdous Rahman Sarker, Farhana Haque, Avinash Sharma, Danai Papakonstantinou, Priyamvada Paudyal, Md Asaduzzaman, Alimuddin Zumla, Najmul Haider

**Author notes:** Corresponding author: Dr Najmul Haider, Lecturer in Epidemiology, School of Life Sciences, Keele University, Staffordshire, United Kingdom, ST5 5BG.

## Abstract

In 2023, Bangladesh experienced its largest and deadliest outbreak of Dengue virus (DENV), reporting the highest-ever recorded annual cases and deaths. We aimed to characterize the geographical transmission of the DENV in Bangladesh. From 1 Jan – 31 Dec 2023, we extracted and analyzed daily data on dengue cases and deaths from the national Management Information System (MIS). We performed a generalized linear mixed model to identify the associations between division-wise daily dengue counts and various geographical and meteorological covariates. The number of Dengue cases reported in 2023 was 1.3 times higher than the total number recorded in the past 23 years (321,179 vs. 244,246), with twice as many deaths than the total fatalities recorded in the past 23 years (1705 vs. 849). Of the 1705 deaths in 2023, 67.4% (n=1015) expired within one day after hospital admission. The divisions southern to Dhaka had a higher dengue incidence/1000 population (2.30 vs. 0.50, p<0,0.01), and higher mean annual temperatures (27.46 vs. 26.54 °C) than the northern divisions. The average daily temperature (IRR: 1.13, 95% CI: 1.11-1.14), urban and rural population ratio of the divisions (IRR: 1.04, 95% CI: 1.03-1.04), showed a positive, and rainfall (IRR: 0.99, 95% CI: 0.98-0.99) showed a negative association with dengue cases in each division. We observed a major geographical shift of Dengue cases from the capital city Dhaka to different districts of Bangladesh with a higher incidence of dengue in the southern division of Bangladesh, influenced by temperature and urbanization.

## Introduction

In 2023, Bangladesh witnessed its most extensive and deadliest dengue outbreak on record, marked by the highest annual tally of cases and fatalities due to dengue virus (DENV) infection [1]. Despite dengue being endemic in Bangladesh, with reported cases annually since 2000, [2] the scale of the outbreak in 2023 is staggering and alarming. Recent years have seen a concerning uptick in dengue cases in Bangladesh, with over 82% of the total cases (n=202,425) and 69% of deaths (n=550) reported in the past five years (2018-2022) [2].

Historically, most of the dengue cases in Bangladesh have been reported in urban areas, with a particular concentration in the capital city of Dhaka [3] except in some years (e.g., 2019) when almost half of the cases were reported from outside Dhaka [4]. Sporadic cases of dengue were documented in Dhaka in the 1960s, preceding the significant outbreak that occurred in 2000 in major cities, including Dhaka, Chattogram, and Khulna [3,5].

Serological studies conducted across the country demonstrated substantial spatial heterogeneity in seropositivity with seroprevalence ranging from as high as 88% in urban Chattogram to as low as 3% in rural Maulvibazar in Sylhet division [6]. In the capital city Dhaka, the seropositivity of DENV ranged from 36 to 85% [6].

*Aedes aegypti,* the primary vector of dengue virus is known for its preference for urban and suburban environments [7]. Several factors contribute to this affinity for urban areas including the presence of artificial containers, human habitation and blood hosts, microclimate in urban areas, and adaptability [7]. On the other hand, *Aedes albopictus*, the second important vector of the dengue virus exhibits a broader habitat range including rural and urban areas [8]. Other factors that affect the spread of the dengue virus are urbanization, population density, rainfall, waste management and watering distribution systems, and temperature [9]. As Bangladesh has recently experienced a country-wide distribution of dengue cases, it is important to understand the factors that affect the geographical distribution of dengue cases in Bangladesh. The objective of this study was to characterize the geographical transmission of dengue virus infection and identify the factors affecting the dispersion of dengue cases in Bangladesh.

## Methods Data Source

We collected the publicly available data on all dengue cases and death records from 1 January to 31 December 2023 from the daily press release of the Management Information System (MIS) of the Ministry of Health and Family Welfare, Bangladesh [10]. The MIS defined dengue cases based on clinical symptoms (including fever and rash) and laboratory tests for IgM or IgG antibodies to DENV and/or nonstructural 1 protein (NS-1) of DENV [11]. MIS collected data from 77 hospitals based in Dhaka city (20 public and 57 private hospitals) and the district hospitals of 63 other districts of the country including the hospitalized patients in tertiary care medical college hospitals [2]. We collected 3-hourly meteorological data on temperature, relative humidity and daily rainfall from the Bangladesh Meteorological Department (BMD) over the period 2000–2023 from the meteorological station located in divisional headquarters including Agargaon, Dhaka (Lat 23.46, Lon 90.23), Chattogram (Lat 22.16, Lon 91.49), Rajshahi (Lat 24.22, Lon 88.42), Rangpur (Lat 25.44, Lon 89.14), Sylhet (Lat 24.54, Lon 91.53), Barisal (Lat 22.45, Lon 90.20), Khulna (Lat 22.47, Lon 89.32), and Mymensingh (Lat 24.43, Lon 90.26). We drew an imaginary east-west line in the middle of Dhaka city to compare the incidence and weather pattern of the southern (Chattogram, Khulna and Barisal) and northern divisions (Rajshahi, Rangpur, Mymensingh, and Sylhet). As the Dhaka division is located centrally in Bangladesh, it was excluded from the southern or northern part.

### Patient data

We further collected anonymous individual patient data including age, sex, address, and hospital stays from MIS. We plotted the age and gender-wise distribution of cases. We also summarized the hospital stays of the death cases. Hospital stays for the survival cases were not available.

### Relative increase of dengue cases by division

We have estimated monthly relative changes in dengue cases in each division. The relative changes (an increase or decrease) of a division of dengue cases for a month were estimated with the formula as shown below

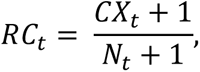

where *RC_t_* is the relative changes of dengue cases in *t* month, *CX_t_* is the number of dengue cases reported in X city, *N_t_* is the total number of cases in Bangladesh in *t* month. To avoid any complication of 0 cases in any city in any month we added 1 dengue case in both numerator and denominator.

### Incidence by district

We calculated the annual cumulative district-wise incidence of dengue cases by taking the cumulative annual number of dengue cases of each district divided by the population of the district shown as - (The total number of dengue cases in a district in 2023) / Total number of populations of that district) *1000. We then generated a map for Bangladesh showing district-wise incidence of dengue cases in 2023. We compared the incidence by divisions (southern vs. northern).

### Statistical Analysis

We compared the dengue cases and deaths of the year 2023 with the previous 23 years (2000- 2022), prepared graphs, plots, and maps, and compared these data with meteorological parameters. We reshaped our dataset by incorporating division-wise outcome variables. We followed the list of districts for each division as shown in the daily dengue situation report shared by MIS [10]. We further collected division-wise population and geographical data from the Statistical Yearbook Bangladesh 2022 published by the Bangladesh Bureau of Statistics [12] including population size, the ratio of rural and urban population (which is a proxy variable for urbanization), and distance from the capital city, Dhaka. Additionally, we calculated population density by dividing the population size by the area of each division district.

We used a generalized linear mixed model (GLMM) with negative binomial distribution to model the division-wise daily dengue counts enhancing modelling flexibility through the inclusion of random effects [13]. We introduced the random effect into the GLMM model to account for the time series effects in the data [12]. The choice of negative binomial distribution allowed us to model response data appropriately with extra variations in the data (overdispersion) [14]. Parameter estimation in GLMMs is challenging due to the integration of random effects in the likelihood function [15]. However, our model results are presented as adjusted incidence rate ratios (IRRs), considering dengue deaths, the urban-rural ratio (as an urbanization proxy), population density, and distance from Dhaka, along with corresponding 95% confidence intervals. The components of the NB-GLMM are given below:

- Distribution: *y*_*ij*_ | *r*_*j*_ ∼ Negative Binomial (*λ*_*ij*_, *φ*),

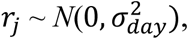

- Linear predictor: *η*_*ij*_ = *β*_0_ + *τ*_*i*_ + *r*_*j*_
- Link function: log (*λ*_*ij*_) = *η*_*ij*_.

where *y*_*ij*_ denotes the number of cases in day *i* on division *j* (*i* = 1, 2, ⋯, 365; *j* = 1, 2, ⋯, 8), *η*_*ij*_ is the linear predictor, *η* is the intercept, *τ*_*i*_ is the fixed effect due to day *i* for the *j*th covariate, and *r*_*j*_ is the random effect due to division *j* [16].

The specific form of our model can be given by

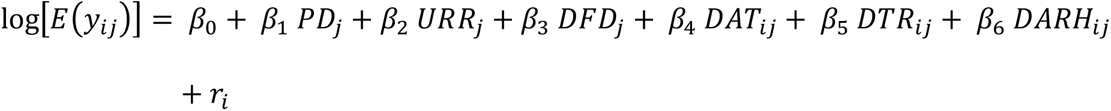

where *PD*_*j*_ is the population density, *URR*_*j*_ is the urban-rural ratio, and *DFD*_*j*_ is the distance from Dahka for the division *j*, *AT*_*ij*_ is the daily average temperature, *DTR*_*ij*_ daily total rainfall, and *DARH*_*ij*_ daily average relative humidity for day *i* and division *j*.

## Results

### Dengue Cases and Deaths

During 2023 (1 January to 31 December), a total of 321,179 dengue cases were reported with 1,705 deaths (case fatality ratio: 0.53%). Between 2000 and 2022, Bangladesh reported a total of 244,246 dengue cases including 849 deaths with a case-fatality ratio of 0.49%. The number of cases reported in 2023 was 1.3 times higher than the number of reported cases in the past 23 years: 2000-2022, (321,179 vs 244,246) and two times more deaths than the number of fatalities recorded in the past 23 years (1,705 vs. 849) in the country **(Fig. 1)**. The number of reported cases and deaths was higher in each month in 2023 compared to the average number of cases or deaths in the corresponding months from 2000 to 2022 **(Fig. 1)**.

**Fig. 1.**
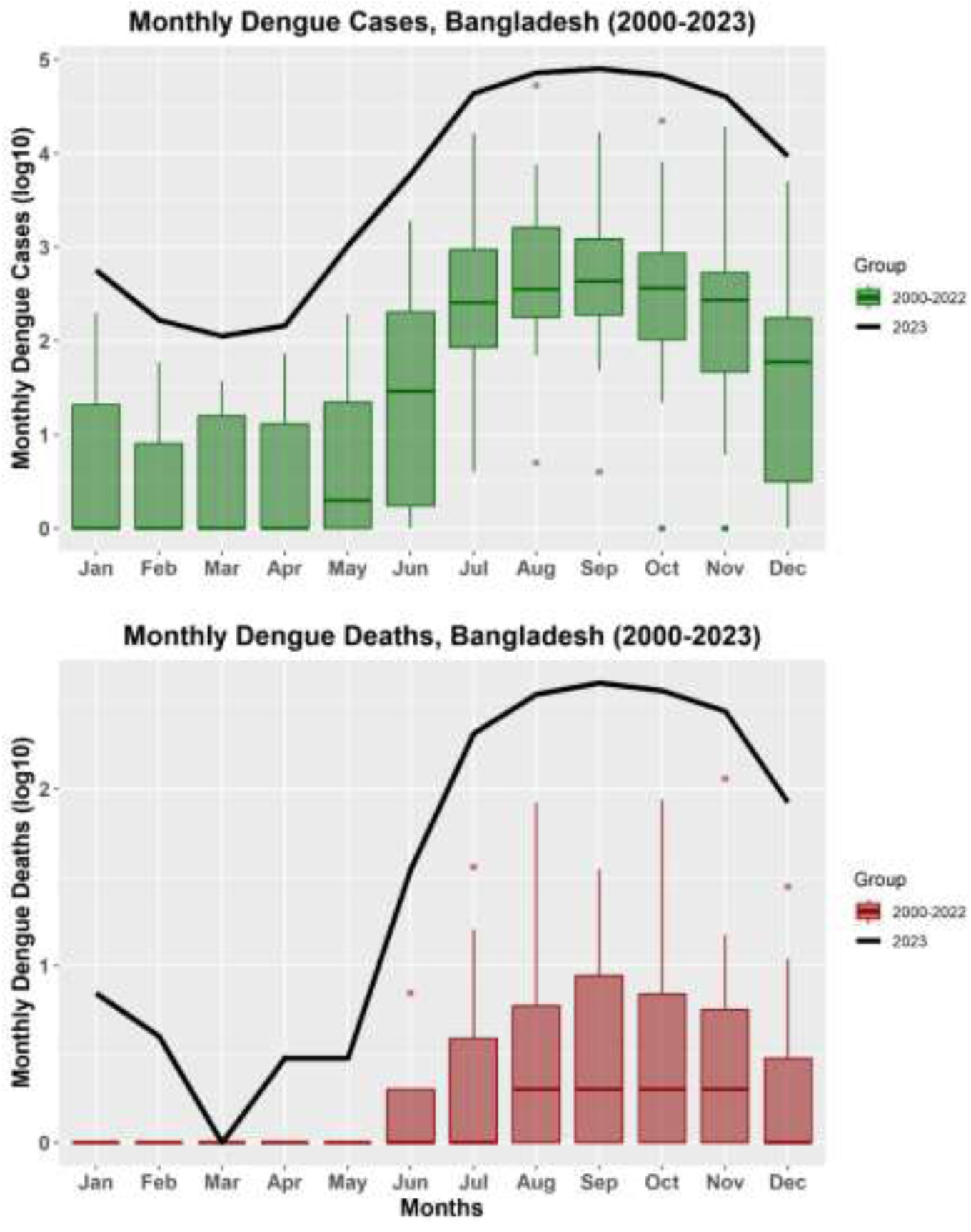
The total number of dengue cases and deaths reported in each month in 2023 vs 2000- 2022 in Bangladesh. Log 10 base is used to display the cases and deaths for the convenience of visualization and comparison.

Among the individuals with dengue cases, 40% were female and 56% were below 30 years of age. A total of 110,008 cases were reported from the capital city of Dhaka including 980 deaths (case-fatality ratio: 0.89%) while 211,171 cases were reported from outside Dhaka including 725 deaths (case-fatality ratio of 0.34%). A higher proportion of cases were detected among young adults of <30 years [55 vs. 45%] but a greater proportion of deaths were detected among older adults of >30 years (68 vs 32%) **(Fig. S1 in the Supplementary material)**. Although males constituted a higher percentage of cases (60 vs 40%), females constituted a greater proportion of deaths (57 vs. 43%) **(Fig. S2 in the Supplementary material).**

Of the 1,705 people who died in 2023, 67.4% (n=1,015) died within one day after hospital admission, with a mean hospital stay of 2.5 days (range: 0-61 days). While considering the first 2 days, the death toll increased to 74.6% (n=1273) or 81.9 % (n=1397) in the first 3 days **(Fig. 2)**.

**Fig. 2.**
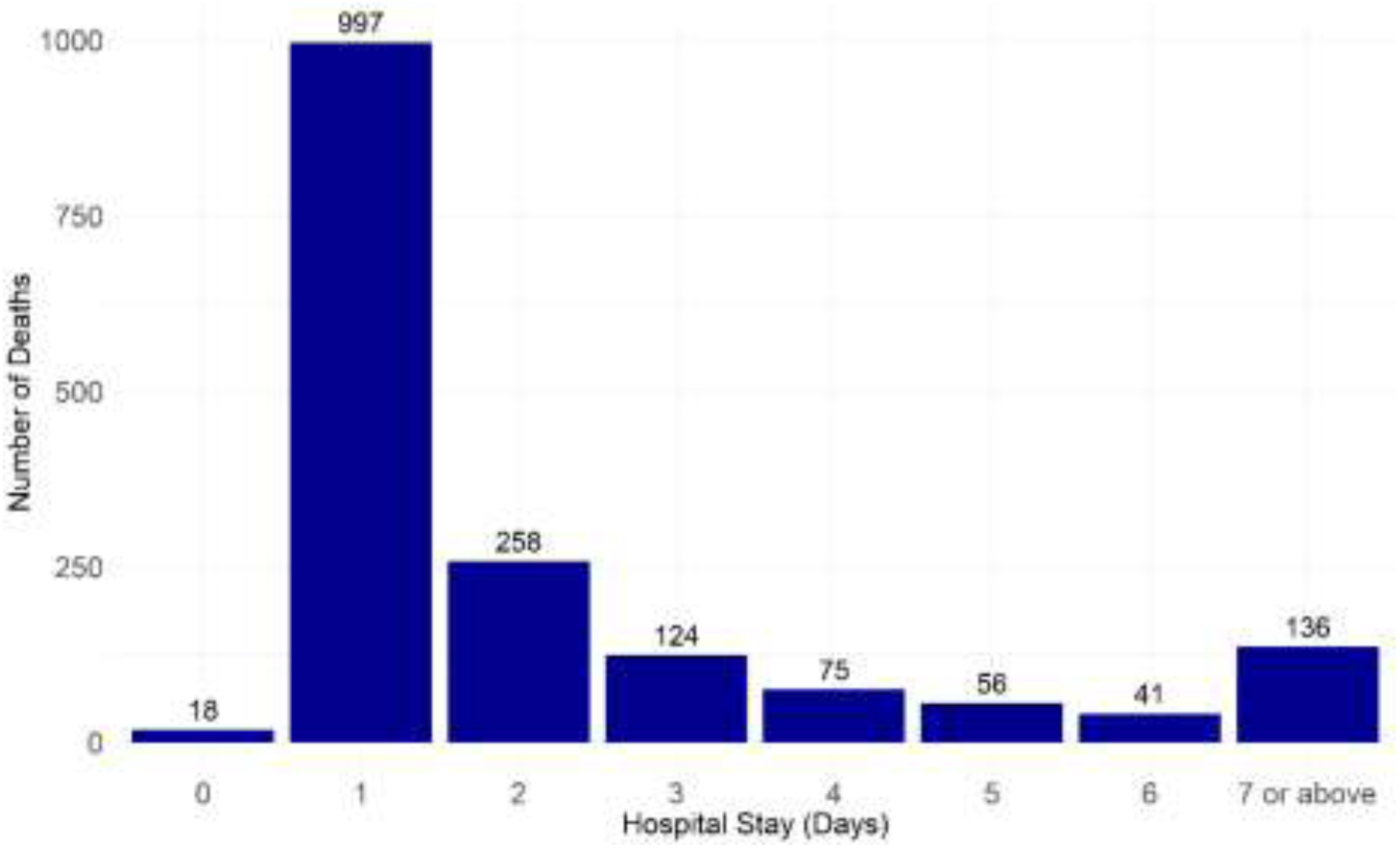
The number of days of hospital stays of 1705 dengue cases in Bangladesh: 1 Jan – 31 Dec 2023. More than 67% (n=1015) of people died within one day of hospital admission.

### Meteorological Data

Bangladesh experienced a higher amount of rainfall in 2023 compared to the average annual rainfall of the period 2000-2022. The average rainfall for the period 2000 to 2022 was 1915.75 mm whereas in 2023 total annual rainfall increased to 2160.70 mm **(Fig. 3)**.

**Fig. 3.**
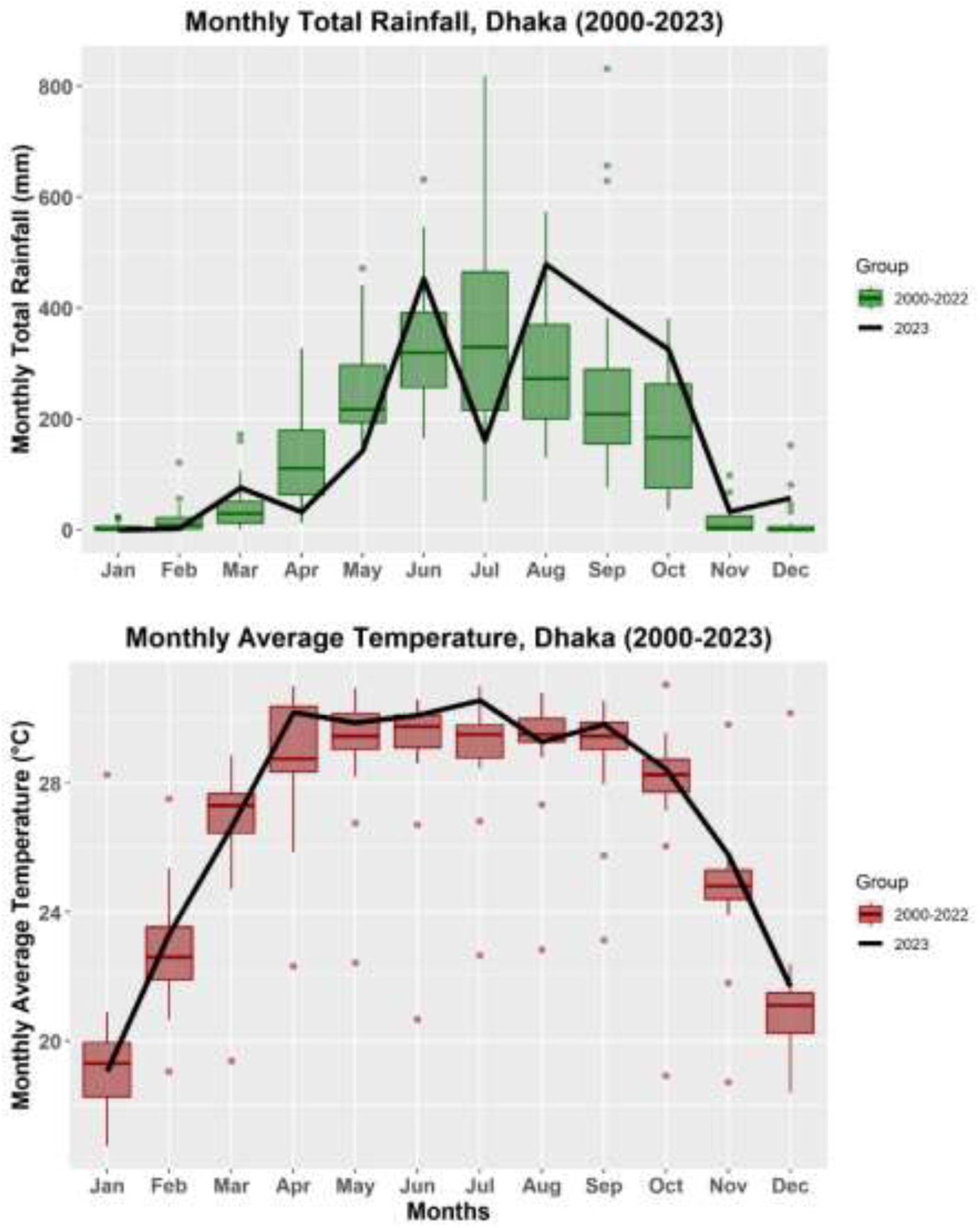
The rainfall (mm) and temperatures (°C) recorded in a weather station in Agargaon, Dhaka, by Bangladesh Meteorological Department, Bangladesh for the period 2000-2022 vs. 2023. Extended monsoon season was observed in 2023 in Bangladesh.

In 2023, rainfall started earlier in the year with 75.8 mm of precipitation in March compared to an average of 45 mm amount of rainfall for the month of the period 2000-2022. There was a similar range of temperature between 2023 and the period 2000-2022 (26.46 °C for the period 2000-2022 vs. 27.06 °C in 2023 **(Fig. 3)**.

### Dengue cases and meteorological data in southern vs. northern Divisions

The divisions southern to Dhaka had a higher dengue incidence compared to the northern division (2.30 vs. 0.50, p<0,0.01) whereas the central Dhaka division had an incidence of 2.90 per thousand population. In 2023, the southern divisions recorded slightly higher annual temperatures (27.46 vs. 26.54 °C) and relative humidity (80.79 vs. 79.08%) than the northern division **(Table 1)**.

**Table 1.** The incidence and case-fatality ratio of dengue and annual temperature and rainfall in 2023 in the Southern and Northern divisions of Bangladesh.

| Parameters | Southern divisions | Northern divisions | p-value | Dhaka division |
| --- | --- | --- | --- | --- |
| Annual mean temperature (°C) in 2023 | 27.46 | 26.54 | <0.01 | 27.07 |
| Annual total rainfall (mm) in 2023 | 2026.5 | 2638.13 | 0.049 | 2160.7 |
| Annual mean relative humidity (%) in 2023 | 80.79 | 79.08 | <0.001 | 70.88 |
| Incidence of dengue (per 1000 population) in 2023 | 2.30 | 0.50 | <0.01 | 2.92 |
| Case-fatality ratio of dengue (%) in 2023 | 0.24 | 0.13 | 0.110 | 0.29 |

### Relative changes in dengue cases in each division

Dhaka city was the primary outbreak site in 2023 and contributed to more than 50% of the total cases up until July and then cases started to increase outside Dhaka, where Dhaka division (excluding Dhaka city) and Chattogram division have been among the prominent sites of the outbreak **(Fig. 4)**. In May, Dhaka city contributed more than 83% of the total cases in the country which dropped to 23.4% in December. The relative changes in dengue cases in different divisions became more evident after July when most divisions started to report an increased percentage of cases and Dhaka city started to report a lower percentage of cases **(Fig. 4).** In November, the Dhaka division (except Dhaka city) reported almost 23% of dengue cases which was the highest percentage of dengue cases for any division in the country, the first record of surpassing the number of cases reported in Dhaka city by any division of the country (**Fig. 4)**.

**Fig. 4.**
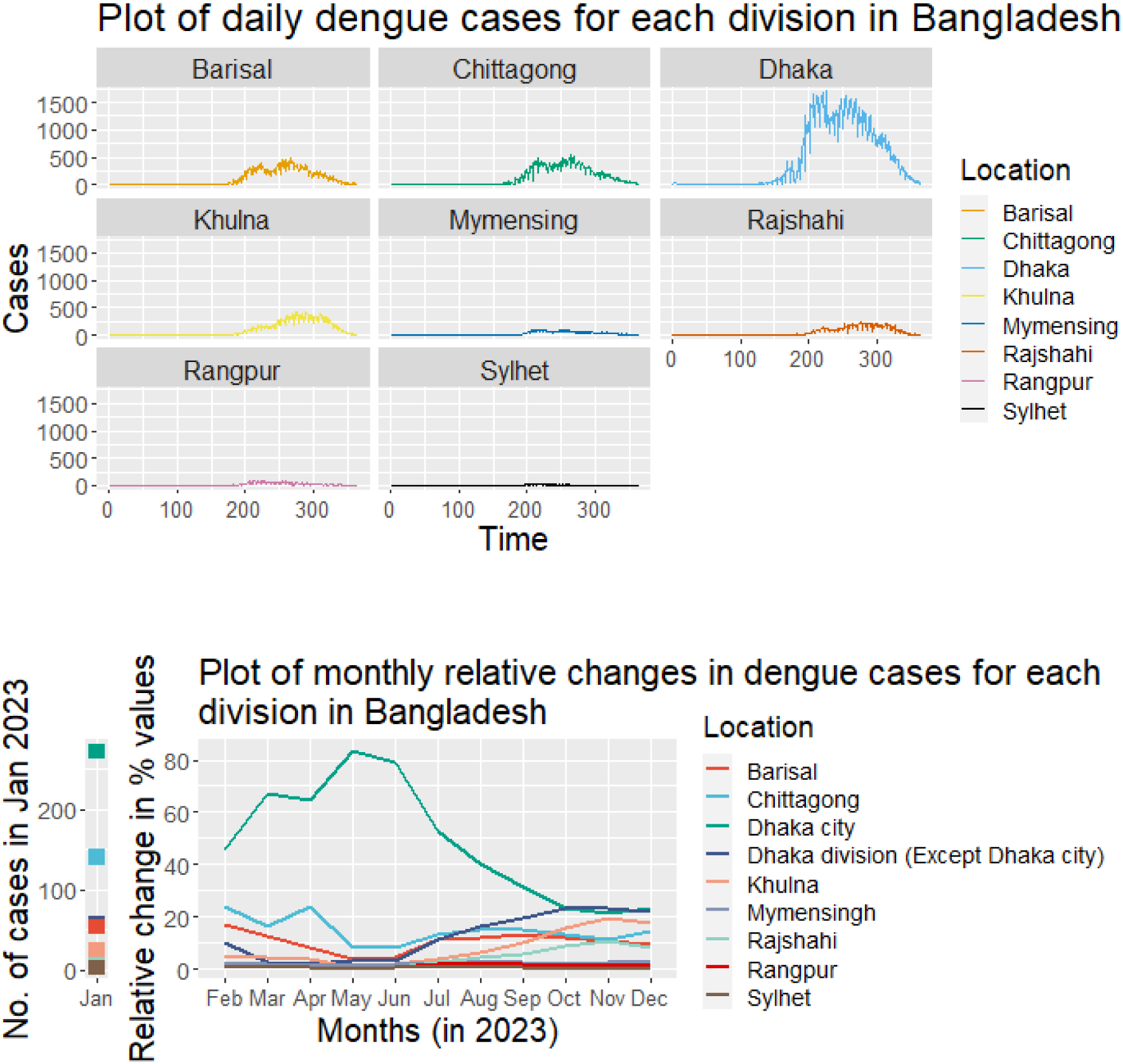
A (Top) The daily number of dengue cases in different divisions of Bangladesh (1 Jan – 31 Dec 2023). B (Bottom). The monthly relative changes of Dengue cases in each division in Bangladesh, 2023 from previous months. Although Dhaka city remains the centre of the outbreak, the percentage of cases has increased outside Dhaka city after July 2023.

The Sylhet division contributed to less than 1% of cases throughout the year. The amount of annual total rainfall recorded in the northern divisions was 2638.13 mm as compared to 2026.50 mm rainfall in the southern divisions (p<0.01). The mean annual temperature recorded in the southern divisions was 26.60 °C as compared to the 25.77 °C temperature of the northern divisions. The temperature of Dhaka division was 27.07 °C and rainfall was 2160.7 mm.

We compared the number of dengue cases in the capital city Dhaka vs. the rest of the country (**Fig. 5)**.

**Fig. 5.**
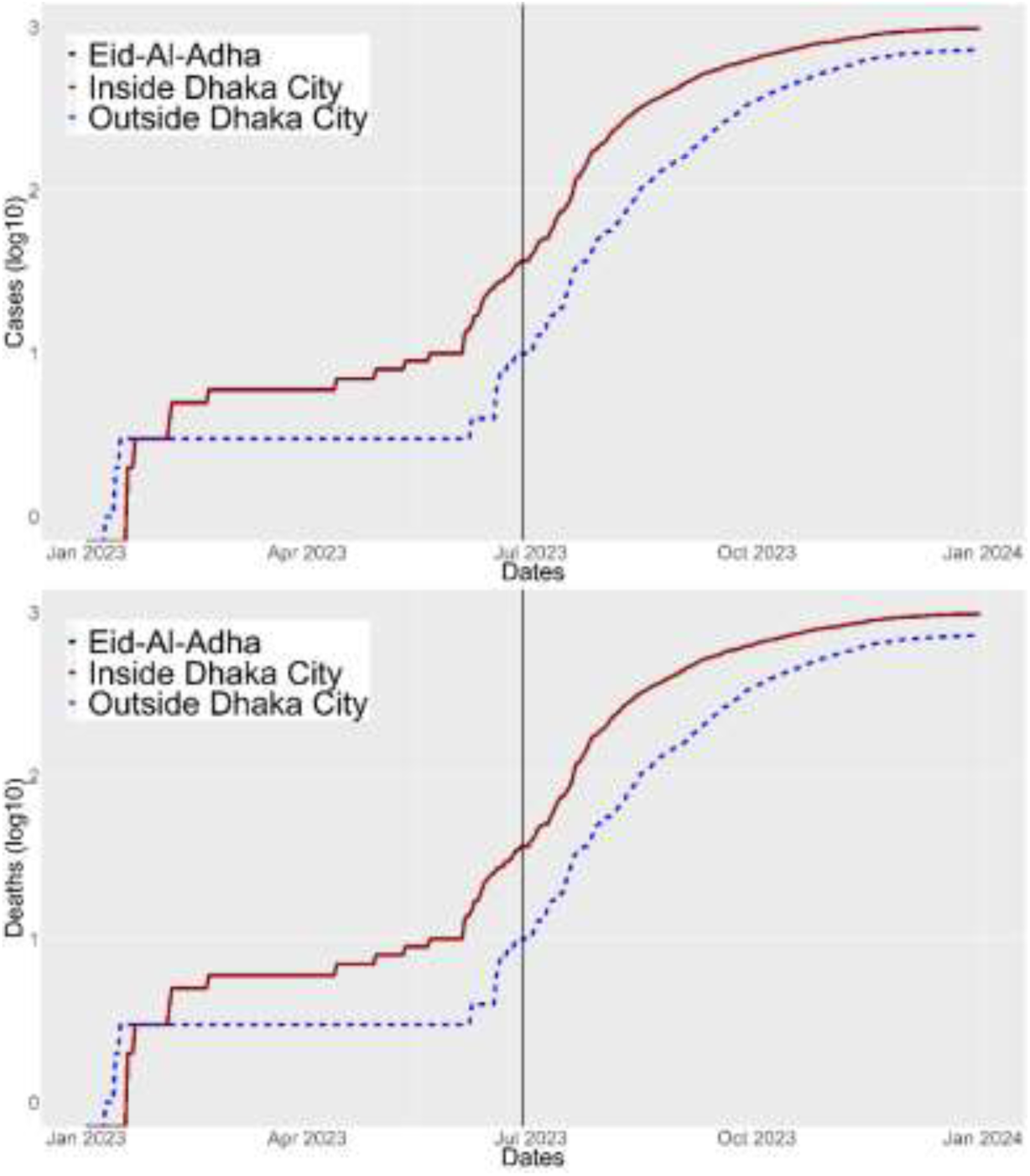
The line graph of dengue virus infection in the capital city Dhaka and outside from 1 January to 31 December 2023. A large number of people from the capital city left Dhaka when Eid-Al-Adha was celebrated on the 28^th^ of June and subsequently, dengue cases started to increase outside Dhaka.

Of 320,835 dengue cases, 207,716 (65%) were reported from outside Dhaka, whereas more than 57.5% (980 of 1705) deaths were recorded in Dhaka. There was a parallel trajectory in both Dhaka city and outside until mid-April. After that, dengue cases started to increase exponentially in the capital city Dhaka which continued up until the end of July 2023, and then the number of cases outside Dhaka surpassed the capital city. Notably, dengue-related deaths were initially higher outside Dhaka City until February, after which an escalation within Dhaka City commenced and persisted till the end of the year.

District-wise, Dhaka district reported the highest number of dengue cases at 113,233, followed by Chattogram (14,200), Barisal (13,603), Manikganj (12,952), and Patuakhali (7,579). On the contrary, the lowest Dengue cases were recorded in Sunamganj (102), Maulvibazar (129), Panchagarh (187), Joypurhat (264), and Lalmonirhat (305). For the number of dengue-related deaths, Dhaka reported the highest death toll at 981, trailed by Barisal (167), Faridpur (138), Chattogram (106), and Khulna (41) districts **(Fig. 6)**.

**Fig. 6.**
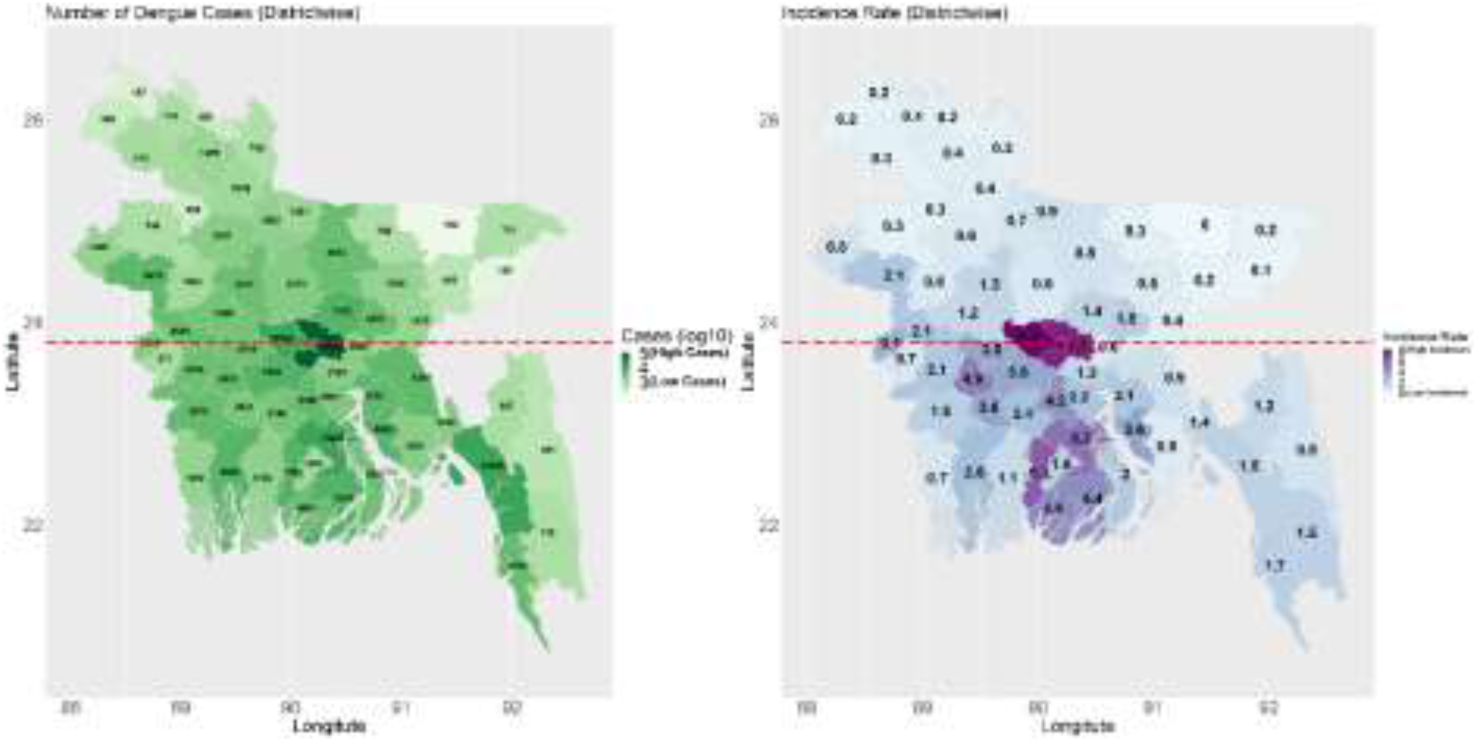
A (Left). The distribution of dengue cases in different districts of Bangladesh, 1^st^ Jan 2023 – 31^st^ Dec 2023. **B (Right)** The incidence of dengue cases in each district in Bangladesh (1^st^ Jan- 31^st^ Dec 2023). The horizontal line in the middle of the country divides the southern and northern divisions.

The southern divisions (Khulna, Barisal, and Chattogram) have a higher mean incidence (2.30 vs. 0.50) and case-fatality ratio (0.24 vs. 0.13) of dengue cases than the northern divisions. The southern division also had a higher annual mean temperature (27.46 vs 26.54 °C) compared to the northern divisions in 2023.

### Correlations

When compared with monthly dengue cases, a positive correlation was observed between population size and the number of dengue cases (*r*=0.44, p=<0.001) and deaths (r=0.43, p=<0.001). A similar association is evident in the relationship between population density and dengue cases (r=0.47, p=<0.001) and deaths (r=0.43, p=<0.001). Conversely, a negative correlation was identified between the distance of each district from Dhaka city and the occurrence of Dengue cases (r=-0.32, p=0.011) **(Fig. S3 in the Supplementary material)**.

In the GLMM, a statistically significant positive association was identified between the dengue cases and daily average temperature (IRR: 1.13, 95% CI: 1.11-1.14), daily average relative humidity of the division (IRR: 1.09, 95% CI: 1.08 – 1.09), urban and rural population ratio (IRR:1.04, 95% CI: 1.03-1.04). Daily total rainfall of the division (IRR: 0.99, 95% CI: 0.98-0.99), showed a significantly negative association between dengue cases. Population density and distance from Dhaka also exhibited weak negative associations **(Table 2)**.

**Table 2.** Factors associated with dengue cases in different divisions using a generalized linear mixed model during 1 Jan 2023 and 31 Dec 2023.

| Variables | Incidence risk ratio (IRR) |  |
| --- | --- | --- |
|  | 95% Confidence Interval | P-value |
| Urban-rural ratio | 1.04 (1.03 – 1.04) | <0.001 |
| Population density | 0.99 (0.99 – 1.00) | 0.056 |
| Distance from Dhaka (capital city) | 0.99 (0.99 – 1.00) | 0.005 |
| Daily average temperature | 1.13 (1.11 – 1.14) | <0.001 |
| Daily total rainfall | 0.99 (0.98 – 0.99) | <0.001 |
| Daily average relative humidity | 1.09 (1.08 – 1.09) | <0.001 |
| **Groups Name** | **Variance** | **Standard Deviation** |
| Location (Intercept) | 0.01652 | 0.1285 |
| **Akaike information criterion (AIC)** | **Bayesian Information Criterion (BIC)** | **Root Mean Square Error (RMSE)** |
| 23720.9 | 23774.8 | 181.804 |
| **Conditional $R^2$** | **Marginal $R^2$** | **Intraclass correlation (ICC)** |
| 0.436 | 0.435 | 0.002 |

## Discussion

In 2023, the world witnessed the first landmark of 6000 annual deaths due to dengue virus infection [17] and Bangladesh reported more than one-fourth of the total global fatalities (n=1,705). In addition to a very high number of dengue cases and deaths, Bangladesh’s 2023 dengue outbreak shows some unique characteristics including i) a widespread distribution of cases nationwide outside Dhaka, ii) a large proportion of deaths (67%) recorded within the first 24 hours of hospital admission, iii) a very high case-fatality ratio of dengue cases within capital City Dhaka, and iv) a higher incidence of dengue cases in the southern divisions of Bangladesh.

Several drivers could have contributed to the largest dengue outbreak in Bangladesh. First, the dengue serotype -2 (DENV-2) reappeared in Bangladesh after 2018 [18]. The absence of the serotype allowed a large proportion of the population to be immunologically naïve as the city experienced more than 4.4% annual growth of population [19]. Second, the outbreak in 2022 was aided by a relatively warmer year and persisting rainfall later in the year which may have collectively facilitated the continued occurrence of dengue cases through January 2023 [18]. Thus, the year 2023 started with as many as 566 cases in January compared to the monthly mean of 126 cases in Bangladesh (2000-2021) [18]. Third, high rainfall in the pre-monsoon season in 2023 (90 mm in March compared to 46.3 mm of the monthly average) allowed increased breeding of mosquitoes leading to an early and consequently large outbreak in the county.

A large majority of deaths (67%) occurred within the first day of hospital admission, suggesting severe disease and/or a considerable delay in seeking medical care. The precise cause of these deaths warrants thorough investigation. Below, we outline two possible explanations for this delay in seeking hospitalization. First, numerous patients likely arrived at the hospital only at the eleventh hour. While primary dengue infection tends to be mild and self-limiting, subsequent infection may escalate to severe forms due to antibody-dependent enhancement and other unknown mechanisms [20]. Distinguishing between primary and subsequent dengue infection is often challenging. Hence, we advocate for the documentation or self-preserving of dengue test results in regions where health data is not recorded systematically. Second, a significant portion of dengue patients (44%) [1] travelled to Dhaka from areas outside the capital city for treatment. These individuals either sought medical attention at a critical stage or were transferred after spending several days admitted to hospitals in districts or sub-districts, with those initial days not being counted as part of their final hospital admission. Many of these patients endangered their lives by undertaking long journeys to Dhaka without access to intravenous fluid during travel. In Bangladesh specialized medical care and management including the facilities for ICU beds are centralized in the country’s capital city Dhaka [1].

Dhaka is one of the most densely populated cities in the world with more than 22 million people living in approximately 300 square Kilometres, with a population density of 23,234 people/Km^2^ [21]. Many people travel to their rural homes during two large religious festivals: Eid-Al-Fitr and Eid-Al-Adha. In 2023, the Eid-Al-Adha was celebrated on 28^th^ June. Up until 28^th^ June 2023, a total of 7862 patients were recorded in the country of which 6014 (76.5%) were recorded in the capital city. More than 15 million people left Dhaka and its surrounding cities to celebrate Eid-Al-Adha with their families in rural Bangladesh [22]. This large population movement probably played a role in spreading the DENV throughout the county. People infected with DENV can remain viraemic (infectious) for a maximum of 12 days [23]. Although *Aedes aegypti*, the key vector of DENV transmission is a city-adapted mosquito, *Aedes albopictus,* is adapted more to rural settings. Earlier studies in Bangladesh reported the presence of *Aedes albopictus* in different parts of Bangladesh [6,24]. In 2023, infected people traveling from Dhaka to rural areas may have spread the virus to the rural areas where the *Aedes albopictus* mosquito maintained the local transmission [1]. In contrast to the idea of an urban disease, dengue might pose a significant threat to rural communities in Bangladesh.

The earlier start of the monsoon in 2023 also coincided with this and may have further influenced the growth of the vector population in rural areas [25]. By 25 July 2023, all 64 districts reported at least one DENV infection in their hospitals. In that specific time, a total of 37, 688 patients were recorded in the country of which 22,349 (59.30%) were recorded in the capital city. The spreading of DENV across the country might have severe consequences for the ongoing outbreak. The rural cycle of DENV transmission is usually led by *Aedes albopictus* and there is some specific difference that makes *Aedes albopictus* a crucial vector for DENV. The *Aedes albopictus* mosquito can bite non-human hosts, tends to bite outdoors, and breeds in tree holes and other natural settings which gives them better plasticity than *Aedes aegypti* [26].

The southern divisions of Bangladesh had a higher incidence and CFR of dengue cases in 2023. Although Bangladesh is a small country there are some differences between the southern and northern parts of Bangladesh as districts in the southern parts observe a higher rate of urbanization and high population density. Also, the divisions in the south of Dhaka had 0.92 °C higher temperature (27.46 vs 26.54 °C, p<0.01) compared to the divisions in the north to Dhaka. Higher temperature has been associated with increased dengue cases because of its impact on the extrinsic incubation period of the virus and the increased biting rate of the mosquitoes [2,27,28]. However, it might be possible that a higher incidence of dengue cases in southern districts is an artifact of economic development in the regions which helped people visit healthcare facilities more frequently than their northern counterparts [29]. Our model also showed that the ratio of urban and rural population which we used as a proxy to indicate urbanization had an increased risk of having more dengue cases. We found a conflicting negative association between rainfall and dengue cases [28], which might be because of higher rainfall in the Sylhet division where the highest amount of precipitation is recorded in Bangladesh. However, the relative humidity was positively associated with increased dengue cases as has been seen elsewhere [28]. Further research is needed to confirm the reason for the higher incidence of dengue cases in southern parts of Bangladesh.

The case-fatality ratio (CFR) observed in 2023 is 10 times higher than the World Health Organization’s (WHO) goal to limit the dengue-related CFR below 0.05% [30]. The CFR of primary DENV infection is generally low with an estimated value of 0.01-0.1%, but the CFR could reach up to 1-4% for secondary or tertiary DENV infection [31]. In the past 23 years, Bangladesh recorded a CFR of 0.34% which is high compared to other countries in the region [2]. In 2023, the CFR is much higher (0.53%) which is inflated by a very high fatality ratio in the capital city Dhaka (0.88%). The high CFR in Dhaka city can be explained as a possible higher rate of secondary or tertiary cases as more than 80% of people in Dhaka city were exposed to any one serotype of DENV in the past [6]. Moderate to severe cases outside of Dhaka city have been referred and travelled to hospitals in Dhaka for better health care, especially for ICU needs. WHO’s situation report reveals that 41% of the death cases were referred to larger cities, especially Dhaka [32]. More than 44% of patients with DENV infection admitted to hospitals in Dhaka city were from outside Dhaka [1]. Also, there is a more regular and organized notification of deaths from Dhaka city as compared to other parts of the country where deaths might be underreported especially when dengue cases are admitted in the private health care facilities. Bangladesh dengue surveillance is only based on selected hospital admissions, which account for approximately 5% of total hospitals in the county, and the patients outside these hospitals as well as private clinics and those not attending any health care settings are not included [1]. Thus, the current passive surveillance system might underreport a substantial number of patients in the denominator of the CFR estimation. However, patients with moderate and severe diseases are likely to be admitted to hospitals, and thus, the deaths are more likely to be notified as compared to the overall infected cases. Thus, it might be worth mentioning that the CFR that we are reporting is more of a CFR for moderate and severe dengue cases, as the denominator might miss a substantial proportion of non-severe dengue cases.

Until the reappearance of DENV-serotype 3 in 2019, the DENV virus was endemic primarily in urban settings, with a large portion of people being exposed to the virus in their lifetime [6]. The distribution of *Aedes aegypti*, which is an urban-dwelling mosquito, probably played a role in such high seroprevalence [6]. This high seroprevalence in large cities, especially in metropolitan Dhaka, created the opportunity for exposure to second, third, or fourth infection with heterogenous serotypes [33]. All four serotypes of the dengue virus have been recorded in Bangladesh at different times since 2000 [18,33]. DENV- Serotype 3 caused a larger outbreak in 2019 and remained a dominant serotype until 2022. DENV-4 reappeared in the year 2022 with co-circulation of DENV-1 and DENV-3. In 2023, DENV-2 became a predominant serotype (62%) along with DENV-3 (29%) and co-infection of DENV-2 and DENV-3 (10%) [34]. Thus, exposure to heterogenous serotypes likely increased the risk of severe dengue infection due to secondary and/or tertiary dengue infection, which has a much higher CFR than the primary infection [23].

As a limitation, the current dengue cases and deaths have been recorded through hospital-based passive surveillance in Bangladesh. The surveillance covers a mere fraction (5%) of the country’s total health healthcare facilities [1]. While we observed significant differences in dengue incidence and CFR between the southern and northern divisions, potential biases linked to the passive surveillance method cannot be ruled out. While improbable, there’s a chance that district health officials in the southern division may have reported more diligently than those in the northern divisions, despite the reporting system being the same throughout the country.

In conclusion, the 2023 dengue outbreak in Bangladesh is marked by an increased occurrence of dengue cases in the southern divisions, with cases spreading geographically from the capital, Dhaka, to other parts of the country, with a notable 65% of cases reported from areas outside Dhaka. The transmission of dengue cases was facilitated by urbanization, as indicated in the proportion of urban vs rural population, and a higher temperature in the southern districts. A large proportion (67%) of deaths were recorded within one day of hospitalization, indicating a late admission of patients with severe disease. A higher proportion of deaths were recorded in the capital city Dhaka, which might be associated with increased secondary or subsequent infection in the city or care seeking of severe dengue cases from all around the country to the capital city hospitals. Bangladesh needs active case and death surveillance, and vector surveillance that incorporates meteorological data and research to identify the causes of increased deaths for improved dengue care. Improved estimation of mild or subclinical cases, their associated risk factors, and temporal trends are essential for effective public health interventions. In contrast to the idea of an urban disease, dengue poses a significant threat to rural communities in Bangladesh.

## Data Availability

We used data that are publicly available in the daily press release of the Ministry of Health and Family Welfare (https://old.dghs.gov.bd/index.php/bd/home/5200-daily-dengue-status-report). There are no identifiable individual-level data, and ethical approval is not required.

## Acknowledgments

We are grateful to the Ministry of Health and Family Welfare (MoHFW) of Bangladesh for publicly sharing the dengue cases and deaths data. We acknowledge the Management Information System of MoHFW for sharing the dengue cases data publicly. NH, and AZ are part of the PANDORA-ID-NET Consortium (EDCTP 373 Reg/Grant RIA2016E-1609) funded by the European and Developing Countries Clinical Trials Partnership (EDCTP2) programme. NH is a member of the International Development Research Centre, Canada’s grant on West African One Health Actions for understanding, preventing, and mitigating outbreaks (109810-001). AZ is a National Institutes of Health Research senior investigator, and a Mahathir Science Award and Pascoal Mocumbi Award laureate.

## Financial Support

There was no funding for this study.

## Conflicts of interest

The authors declare no conflict of interest.

## Autho’s Contributions

Conceptualization: NH, MA, and MNH. Data curation: MNH, SAAA, MA, MR. Formal Analysis: MNH, NH, MA. Writing original draft: NH, MNH. Supervision: AZ Writing, review, and editing: MR, MU, SAAA, KMR, KKP, MFRS, FH, AS, DP, PP, MA, AZ

## Notes

### Competing Interest Statement

The authors have declared no competing interest.

### Funding Statement

This study did not receive any funding

