## Supplementary Material for "Shifting Geographical Transmission Patterns: Characterizing the 2023 Fatal Dengue Outbreak in Bangladesh"

**Fig. S1. The age structure of dengue cases during 1 Jan-31 Dec 2023 in Bangladesh.** A higher proportion of cases were detected among young adults (<30 years) [55% vs. 45%] but a greater proportion of deaths were detected among older adults (>30 years) (68% vs. 32%).

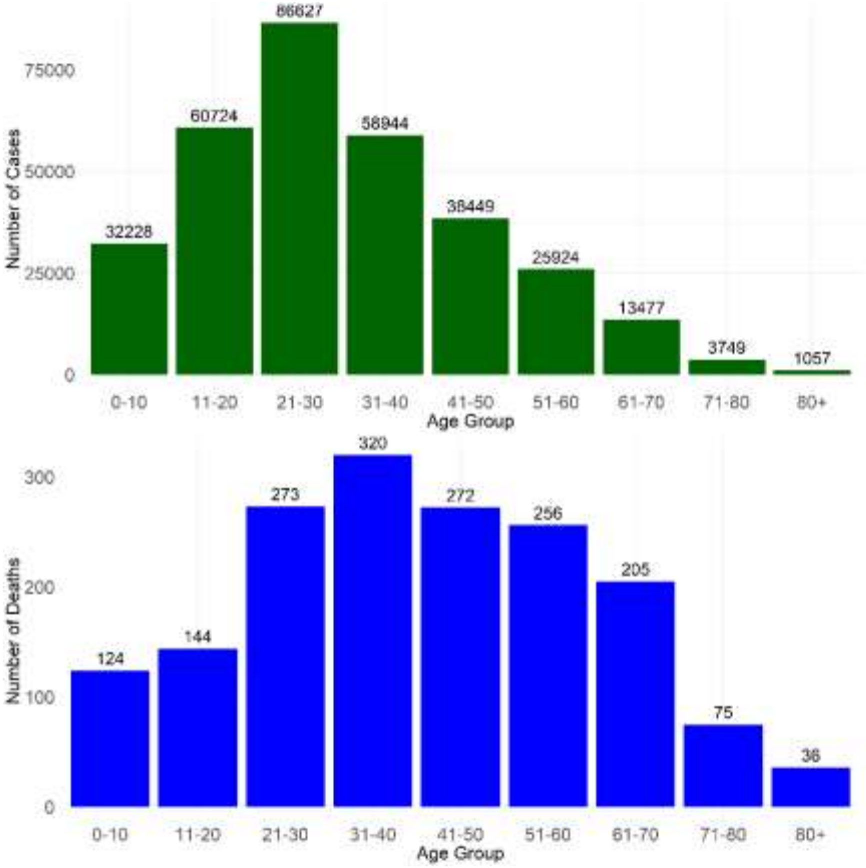

**Fig. S2. The comparison of the proportion of dengue cases and deaths in 2023 in Bangladesh by gender.** Although Males constitute a higher percentage of cases, females constitute a greater proportion of deaths.

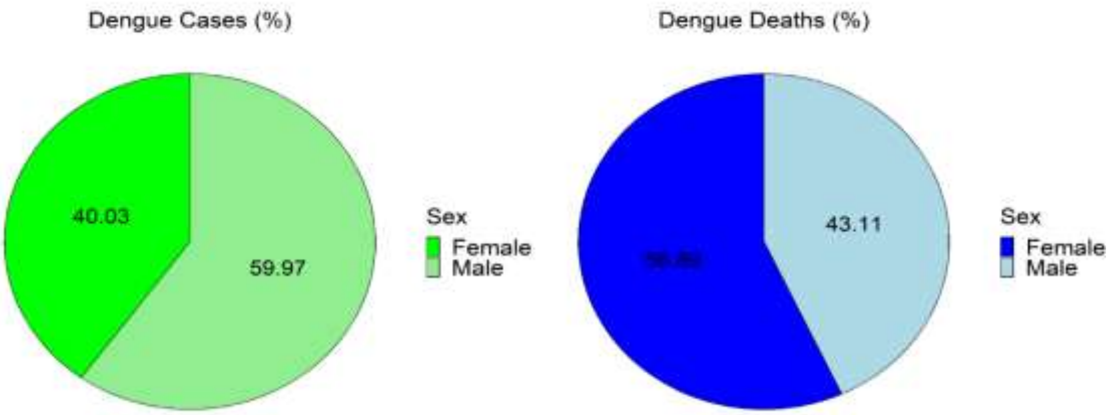

**Fig. S3. The correlation coefficient of dengue cases and deaths in different districts and their population size, population density, and distance from Dhaka city.** A positive correlation exists with the population density of the district and a negative correlation exists with the distance from the capital city Dhaka.

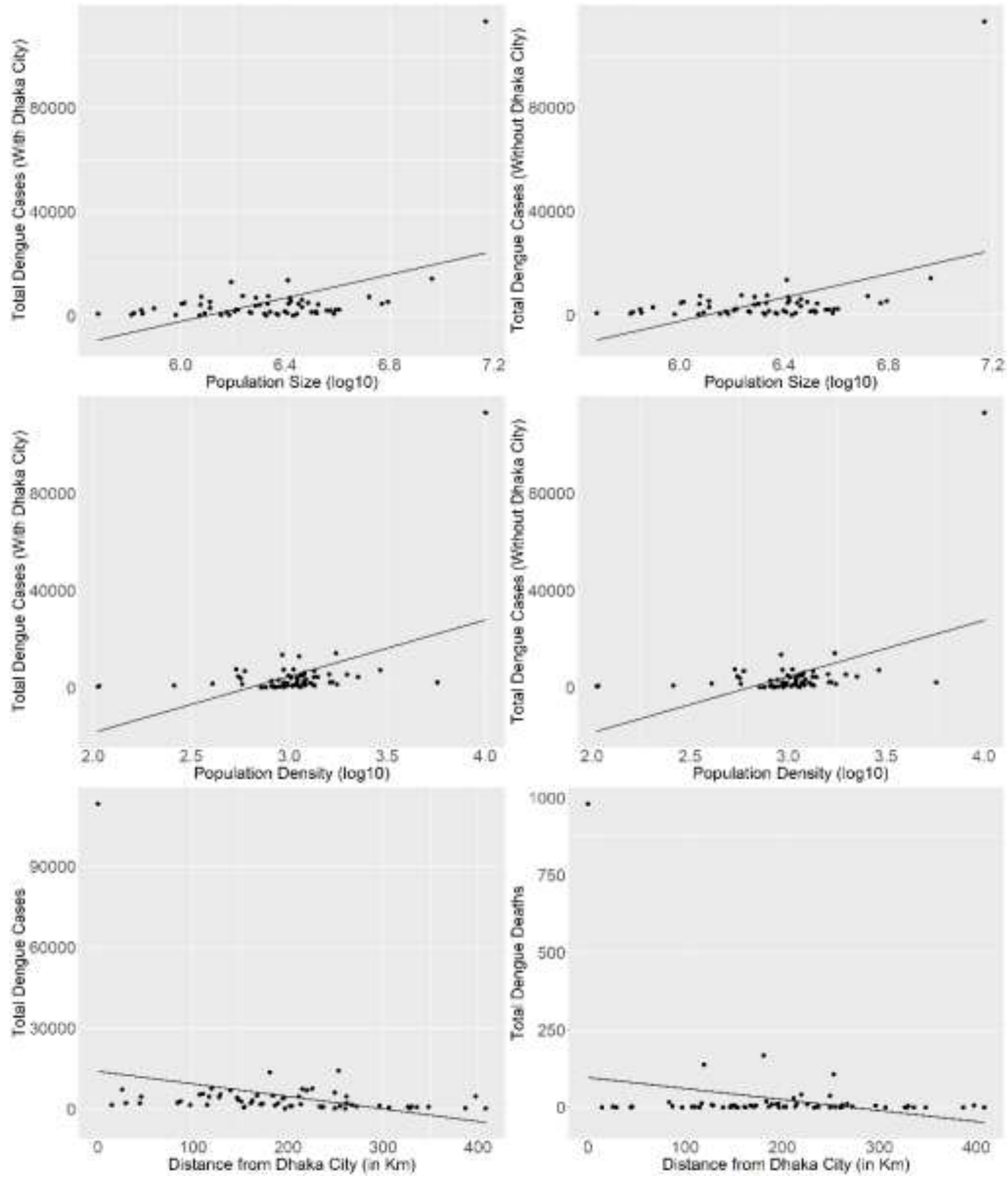
